# Evaluating growth pattern and assessing future scenario of COVID-19 epidemic of India

**DOI:** 10.1101/2020.05.02.20087544

**Authors:** Nandan L. Patil, Lakshmi R. Gangavati

## Abstract

COVID-19 the modern pandemic has spread across the world at a rapid pace. SARS-CoV 2 is highly transmissible and the rate of infection is exponential for heavily infected countries. Asymptotic carriers and longer incubation period have been key towards such a large-scale distribution of disease. Data released by official authorities on COVID-19 cases is significantly affected by various factors such as size of sample, incubation period of disease and time taken to test the sample. These factors mask the useful pattern (signal) of disease spread. Thus, an ingenious method to group data into cycles of five and seven days, for studying pattern of disease spread is undertaken. Occurrence of recurrent peaks as indicated by Adjusted Rate of infection per day indicated the spread of disease has been non-uniform. Currently, India is yet to reach the critical point (peak of epidemic) with adjusted daily cases more than 1000. Increasing testing capacity along with random sampling and sample pooling can help in preventing formation of these peaks in future. The proposed method helps in assessing the current state and for predicting future scenarios epidemics.

## 1. Introduction

Novel coronavirus has swept the world at rapid pace with more than 3 million cases across 185 countries since its first report on November, 2019 in Wuhan, China. The disease was termed as Public Health Emergency by World Health Organization (WHO)on January 30, 2020 and further on 11 March was announced as pandemic. The pandemic has resulted in huge economic cost, loss of jobs and resulting in largest global recession since The Great Depression (Gopinath, 2020). The International Virus Classification Commission (ICTV) classified this novel coronavirus as Severe Acute Respiratory Syndrome Coronavirus 2 (SARS-CoV-2) on February 11, 2020 and on the same day WHO named the disease as COVID-19. Virus is transmitted mainly through respiratory droplets and has high transmissibility rate but low mortality unlike Middle Easter Respiratory Syndrome (MERS) and SARS-CoV 1 which have high mortality rate. The mean incubation period of disease is6.6 with range of 2.1 to 11.1 days(Backer et al., 2020, Huang et al., 2020). COVID-19 symptoms include aches and pains, nasal congestion, runny nose, sore throat or diarrhea and needs technical tests such as RT-PCR, Anti body testing or CRISPAR (Liu et al., 2020) to detect the virus. Many cases are reported were no symptoms are visible but the person is tested positive for the virus. These asymptotic people have been key for such a large scale spread of virus.

India reported its first case on 30 January, 2020 from Southern state of Kerala, who was a returning student from Wuhan, China. Only three cases were reported up till 1 March and since then the cases have increased exponentially and as of April 23, 27000 cases have to identified with Maharashtra recording highest positive cases. The virus has entered the country through foreign travelers coming mainly from China, Europe US and Gulf countries. Foreign flights were stopped on 22 March and nation-wide lockdown was announced on 24 March for 21 days i.e. up till 14 April which was further extended to 3 May. But large population of migrant workers had returned to their native place during initial days of lockdown. Currently, in India, asymptotic people with international travel history and all contacts of laboratory confirmed positive cases are being tested for the virus mainly through RT-PCR. As of now there have been no effective drugs or vaccine against the disease, thus identifying positive cases and isolating/treating them is effective option.

Compartmental models namely SIR (Susceptible, Infectious and Recovered) models and their modification (Giordano et al., 2020, Wu et al., 2020, Prem et al., 2020) are widely used in analyzing the epidemiological data. Furthermore, statistical models such as ARIMA and neural networks (Wei et al., 2016, Anne and Jeeva, 2020, Ceylan, 2020) are used in forecasting the COVID-19 cases. These models become complex when a high number of parameters are involved. Thus, here we formulate a simple statistical method to study the growth of epidemics and predict future scenarios based on only the positive cases reported.

## 2. Methodology

Data of positive cases for COVID-19 epidemic was collected from Johns Hopkins University Center for Systems Science and Engineering (JHU CSSE, 2020) starting from 22 February up to 23 April.

Let the first day *i.e*. February 22 be d_1,_ February 23d_2_ and so on with d_n_ being n^th^ day from the first day.

cc_1_, cc_2_, cc_3_, ………cc_n_ is number of cumulative cases (cc) for the d_1_, d_2_, d_3_, d_n_ day and

dc_1_, dc_2_, dc_3_, ………dc_n_ is number of daily cases (dc) for d_1_, d_2_, d_3_, d_n_ day.

i. Estimating Unadjusted Rate of Infection*(UR*_*d*_):

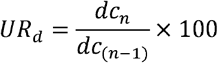

where, *UR*_*d*_ is **U**nadjusted **R**ate of Infection for d^th^ day estimated from raw data, *dc*_*n*_ is number of positive cases for n^th^day, *dc*_(*n*-1)_ is number of positive cases for (n-1)^th^day Cumulative cases (cc) or daily cases (dc) reported by official authorities is a mixture of both useful signal and noise. A clear pattern of epidemic growth cannot be ascertained from the raw data itself. Thus, in order to obtain a clear picture of epidemic growth and to minimize the noise (sampling effect) in the data we group the data based on five day and seven day cycle to estimate new coefficients *R*_*cmi*_ and *Adc*_*n*_as described below. Let ‘*c*’ be the number of days in a given cycle, then number of possible combinations the data can be grouped is equal to ‘*m*’ which are repeated over ‘*i*’ intervals where ‘*m*’ is equal to ‘*c*’. These combinations are referred to as Models from henceforth. In the current study we group the data in cycle of five days (5D) and seven days (7D).
ii. Total positive cases for m^th^ model during i^th^ interval for cycle of seven days (c=7) is calculated as follows,

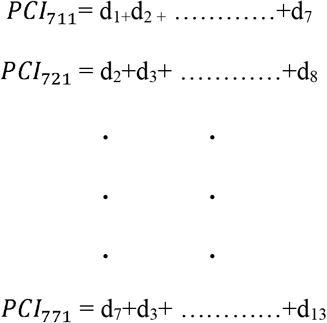

For ‘*c*’ day cycle,

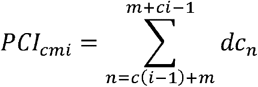

where, c = 1,2, 3…, c and m = 1,2, 3……m and i = 1,2, 3…… *PCI*_*cmi*_ is **P**ositive **C**ases of m^th^ model during i^th^ **I**nterval for ‘*c*’ day cycle *dc*_*n*_ is number of **d**aily **c**ases for n^th^ day
iii. Estimating Rate of infection per ‘*c*’ day cycle (*R*_*cmi*_):

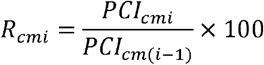

where, *R*_*cmi*_ is **R**ate of **I**nfection for i^th^ interval of m^th^ model for ‘*c*’ day cycle *PCI*_*cmi*_is Positive Cases of m^th^ model during i^th^ interval for ‘*c*’ day cycle *PCI*_*cmi*(*i*−1)_is Positive Cases of m^th^ model during (i-1)^th^ interval for ‘*c*’ day cycle
iv. Estimating the Adjusted Rate of infection per day (*AR*_*cn*_): The **A**djusted **R**ate of **I**nfection of n^th^ day for ‘*c*’ day cycle (*AR*_*cn*_) is calculated as,

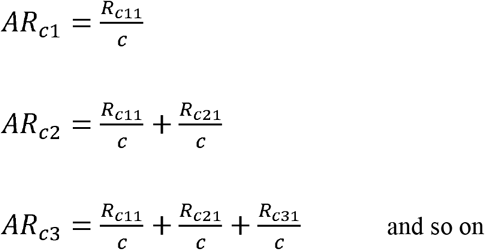
v. Calculation of Adjusted daily cases (*Adc*_*n*_): The **A**djusted **d**aily **c**ases for ‘*c*’ day cycle for n^th^ day (*Adc*_*n*_) is calculated as,

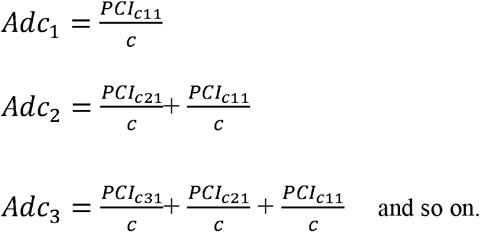

The unadjusted daily cases (*UDc*_*n*_) is defined as daily cases calculated from official record i.e. dc_1_, dc_2_, dc_3_ ……

### 2.1 Outliers

If the magnitude of *PCI*_*cm*(*i*-1)_ is less than 10 it leads to greater *R*_*cmi*_which effects the *AR*_*cn*_and masks pattern in the growth of epidemic. Thus, if *PCI*_*cm*(*i*-1)_< 10, then respective *R*_*cmi*_ are removed from analysis.

### 2.2 Critical Point

Critical point is defined as a stage where the *R*_*cmi*_ or adjusted daily cases (*AR*_*cn*_) are equal to 100 and 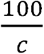, respectively. In other words, critical point refers to period past the peak of the epidemic curve. The *AR*_*cn*_ critical point for 5D and 7D cycle is 25 and 14.29, respectively.

### 2.3 Classifying the growth of epidemic period

Based on the critical point, the epidemic period can be classified into two phases,

a. **Growth phase**: It is the initial phase were epidemic has not yet reached the critical point. During this period of epidemic during which the number of daily cases is continuously rising and epidemic is yet to reach its peak stage.
b. **Safe phase**: It is the phase after passing the critical point or peak of epidemic when the daily cases are continuously decreasing.

### 2.4 Prediction of *AR*_*cn*_ values

ARIMA is a statistical method to forecast time series data which was proposed by Box and Jenkins in 1970. These models have been commonly used to forecast COVID-19 positive cases (Benvenuto et. al., 2020, Ceylan, 2020, Anne and Jeeva, 2020). ARIMA model was used to forecast the *AR*_*cn*_ value of 5D and 7D cycle. The three best fitting parameters of ARIMA model p (order of autoregression), d (degree of difference) and q (order of moving average) were (2, 0, 0) and (3, 2, 1) for 5D and 7D cycle selected based on root mean square error (RMSE). Furthermore, univariate Neural Network Models (NNM) were used in predicting the *AR*_*cn*_ of 5D and 7D cycle by using *forecast* package in R.

## 3. Result and Discussion

COVID-19 pandemic has caused large scale disruption of normal life with nationwide lockdown being imposed to control spread of virus. We have analyzed the data in a novel way to understand growth of virus and anticipate future conditions. Unadjusted rate of infection per day (*UR*_*d*_) plot (Figure 1) estimated from raw data shows continuous fluctuation throughout the epidemic period. No growth pattern of disease can be delineated from the plot. The reason for such fluctuation is that raw data is a mixture of both useful signal which represents growth of epidemic along with redundant noises which mask the signal. The noise in the data arises due to various reasons such as sampling size, time taken to test the sample, time taken to trace the contacts and incubation period for the patient. Thus, making it impossible to assess present situation and predict future conditions using *UR*_*d*_plots.

**Figure 1.**
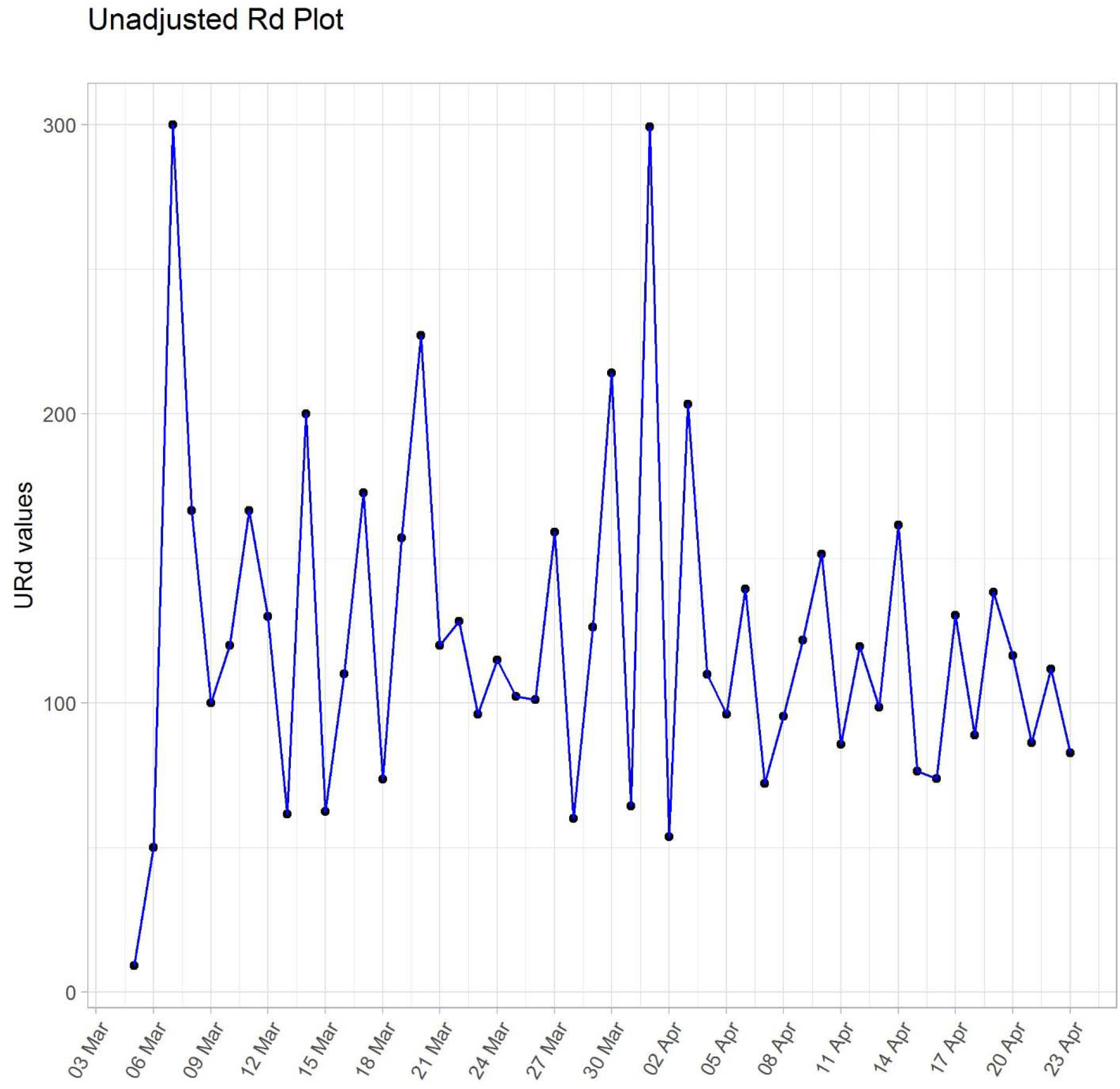
Unadjusted *AR*_*cn*_ from 5 March to 23 April.

### 3.1 Analyzing Adjusted Rate of Infection per day (*AR*_*cn*_ )

In order to reduce the noise embedded in raw data, we grouped raw data into 7D and 5D cycle and estimated the adjusted rate of infection per day. *AR*_*cn*_ plot (Fig. 2 and Fig. 3) for 7D and 5D cycle reveals the pattern of spread of Epidemic. The Epidemic period can be classified into four and three phases (Table 1) as per *AR*_*cn*_ of 5D and 7D cycle respectively. Phase 1 (5D) was from5 March to 16 March, with peak on 14 March and Phase 2 (5D) from 17 March to 28 March with peak on 21 March. The first two phases of 5D combine to form Phase 1 of 7D cycle which was from 5 March to 29 March with peak on 22 March. During these initial stages of epidemic, the virus is difficult to detect due to high incubation period, asymptotic carriers and technical tests to detect the virus. Thus, the virus spreads can spread unnoticed if proper preventive measures are not undertaken. With increase in testing capacity a greater number of COVID-19 cases are detected in short time leading to formation of peak on 22 April.

**Table 1.** List of Phases as delineated by *AR*_*cn*_ of 5D and 7D cycle for India.

| 5D cycle |  |  |  |  | 7D cycle |  |  |  |  |
| --- | --- | --- | --- | --- | --- | --- | --- | --- | --- |
|  | Start | End | Peak | Total cases |  | Start | End | Peak | Total cases |
| **Phase 1** | 5 March | 16 March | 4 March | 90 | **Phase 1** | 5 March | 29 March | 22 March | 996 |
| **Phase 2** | 17 March | 28 March | 21 March | 800 |  |  |  |  |  |
| **Phase 3** | 29 March | 9 April | 2 April | 4947 | **Phase 2** | 30 March | 13 April | 3 April | 8328 |
| **Phase 4** | 10 April | - | 13 April | 15835* | **Phase 3** | 14 April | - | - | 12348* |

\* as of 23 April

**Figure 2.**
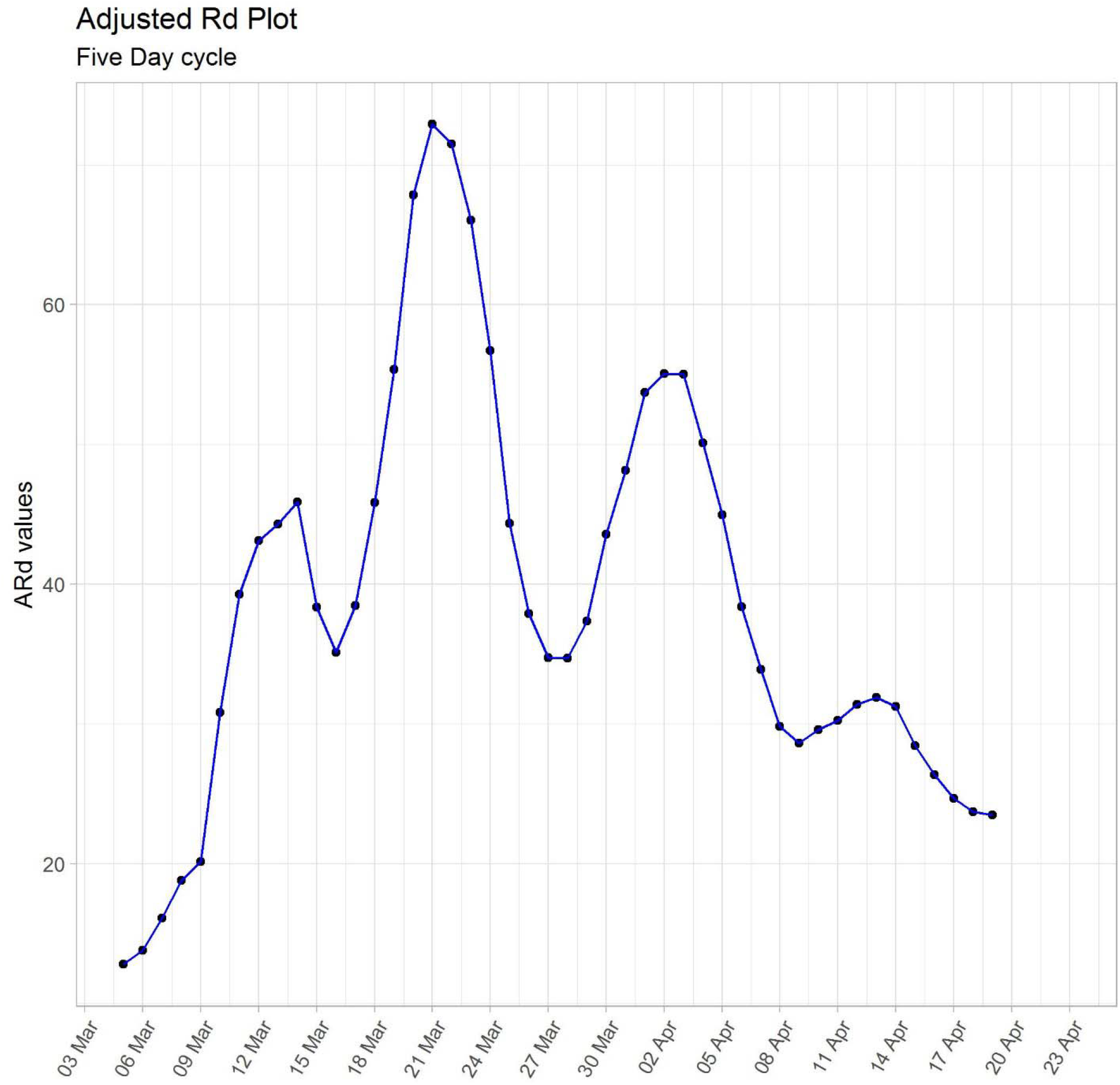
*AR*_*cn*_ plot for five day (5D) cycle from 5 March to 19 March.

**Figure 3.**
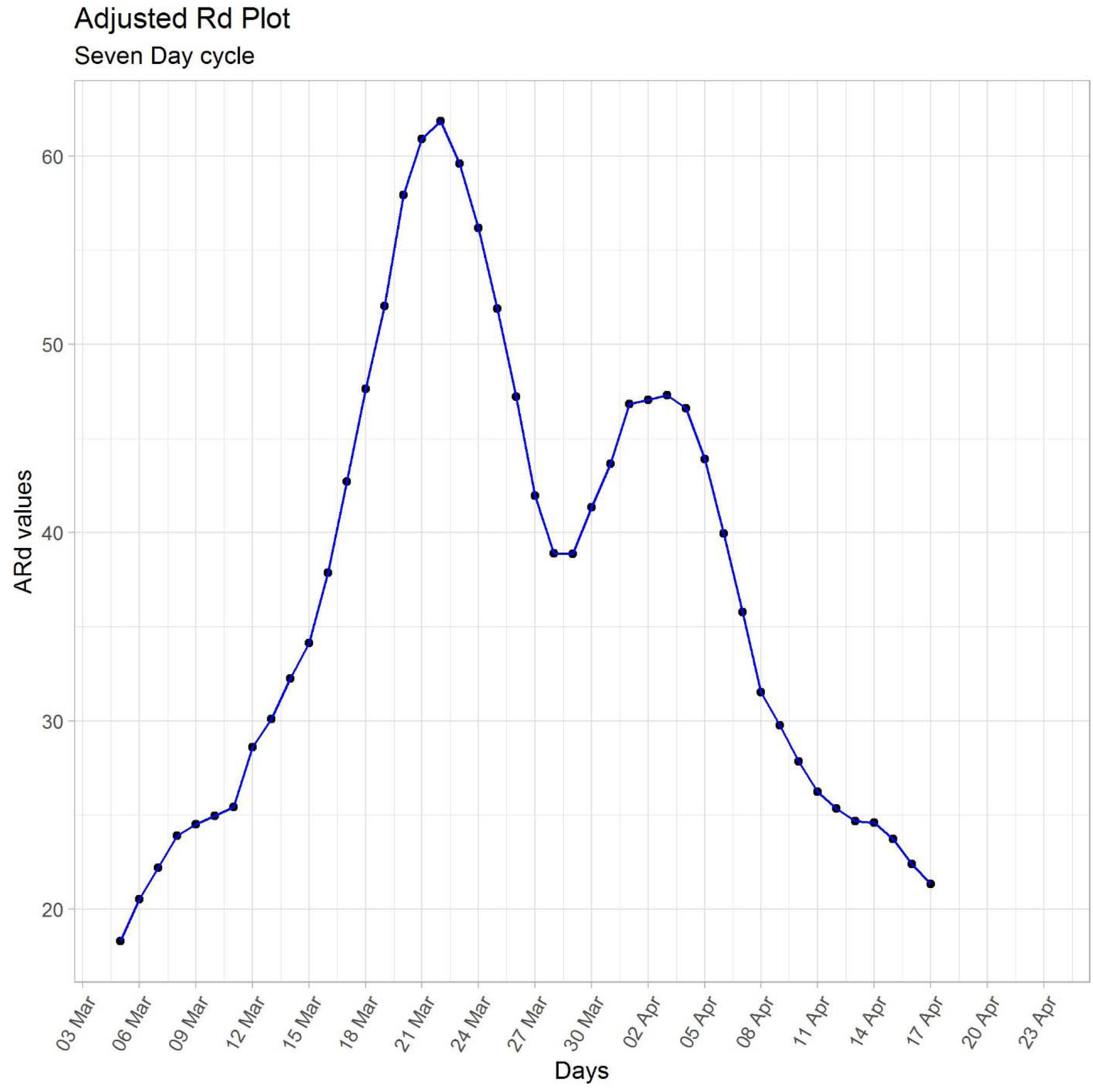
*AR*_*cn*_ plot for seven day (7D) cycle from 5 March to 17 March.

Phase 3 (5D) from 29 March to 8 April with peak on 2 April and Phase 2 (7D) 30 March to 13 April with peak on 3 April can be attributed to Tablighi Jamaat congregation which was held in New Delhi during mid-March. Many of the attendees have been tested positive. Finally, we observe another peak on 13 April under Phase 4 (5D) starting from 10 April. In a *AR*_*cn*_ plot the new peaks formed are more severe than that of early ones. Meaning peak of Phase 4 (5D) is more severe than peak of Phase 3 (5D) and Phase 2 (7D) which is further more severe than peak of Phase 1 (7D) or Phase 2 (5D). The 5D cycle detected a greater number of peaks than 7D cycle as it is more sensitive to sudden increase in cases within short period of time. The recurrent peaks which we observe in *AR*_*cn*_ plots may be attributed to limited coverage of testing facility which allows the virus to spread through asymptotic carriers.

### 3.2 Impact of Nationwide lockdown

Nationwide shutdown started from 24 March and is scheduled to be lifted on May 3. As seen from the *AR*_*cn*_ plot the rate of new infection starts to decrease after its peak on March 22. The nationwide lockdown has significantly slowed the spread of virus, but it should be followed by wide scale testing in order to identify the infected people. Delay in action or lack of testing would lead to increase in *AR*_*cn*_ or under more severe conditions new peaks are formed.

### 3.3 Analyzing *R*_*cmi*_**plots**

*R*_*cmi*_ represents change in rate of infection for a given days of cycle. When the spread is constant the *R*_*cmi*_ of all models for any days of cycle start converging and continue in same direction. But when growth is non-uniform the models start bisecting each other.

For seven-day cycle *R*_*cmi*_ plot (Fig. 4) highest value is 521 of Model 7for interval March 20 to March 26 which coincides with peak of Phase 1 (7D). The models are continuously bisecting each other indicating the spread or detection of virus has not been uniform. Similarly, *R*_*cmi*_plot of five-day cycle indicates same trend of bisecting models with highest value of 455.263 for interval March 20 to March 24 coinciding with peak of Phase 2 (5D). As seen from the analysis of other countries (not shown) the Models tend to converge (or are close to each other) before reaching the critical point.

**Figure 4.**
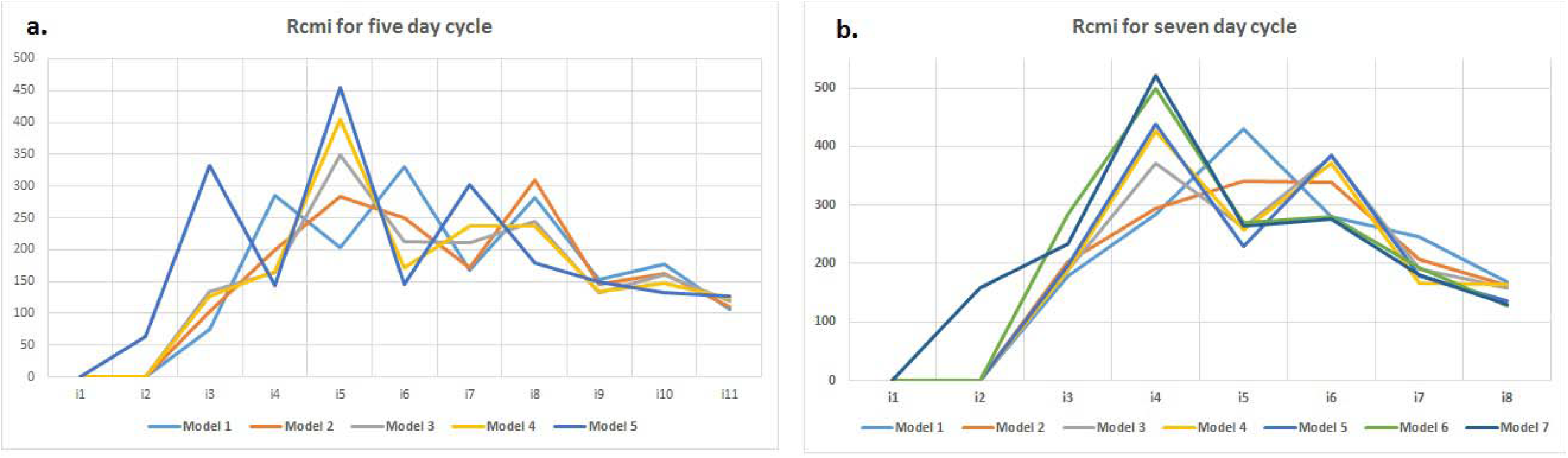
**a)** plots for five day cycle (5D) across 11 intervals, **b)** plot for seven days cycle (7D) across 8 intervals

### 3.4 Adjusted daily cases (*Adc*_*n*_ )

*Adc*_*n*_ accommodates the fluctuation in the original data to produce a smooth curve representing increase in daily cases. India passed *Adc*_*n*_ (Fig. 5) of 1,000 on 14 April and as of 18 April, with current *Adc*_*n*_ being 1173 and 1164 as calculated from 5D and 7D cycle respectively. *Adc*_*n*_ is continuously increasing during growth phase whereas starts decreasing once in safe phase. Thus, delay in reaching the critical point enhances the severity of Epidemic.

**Figure 5.**
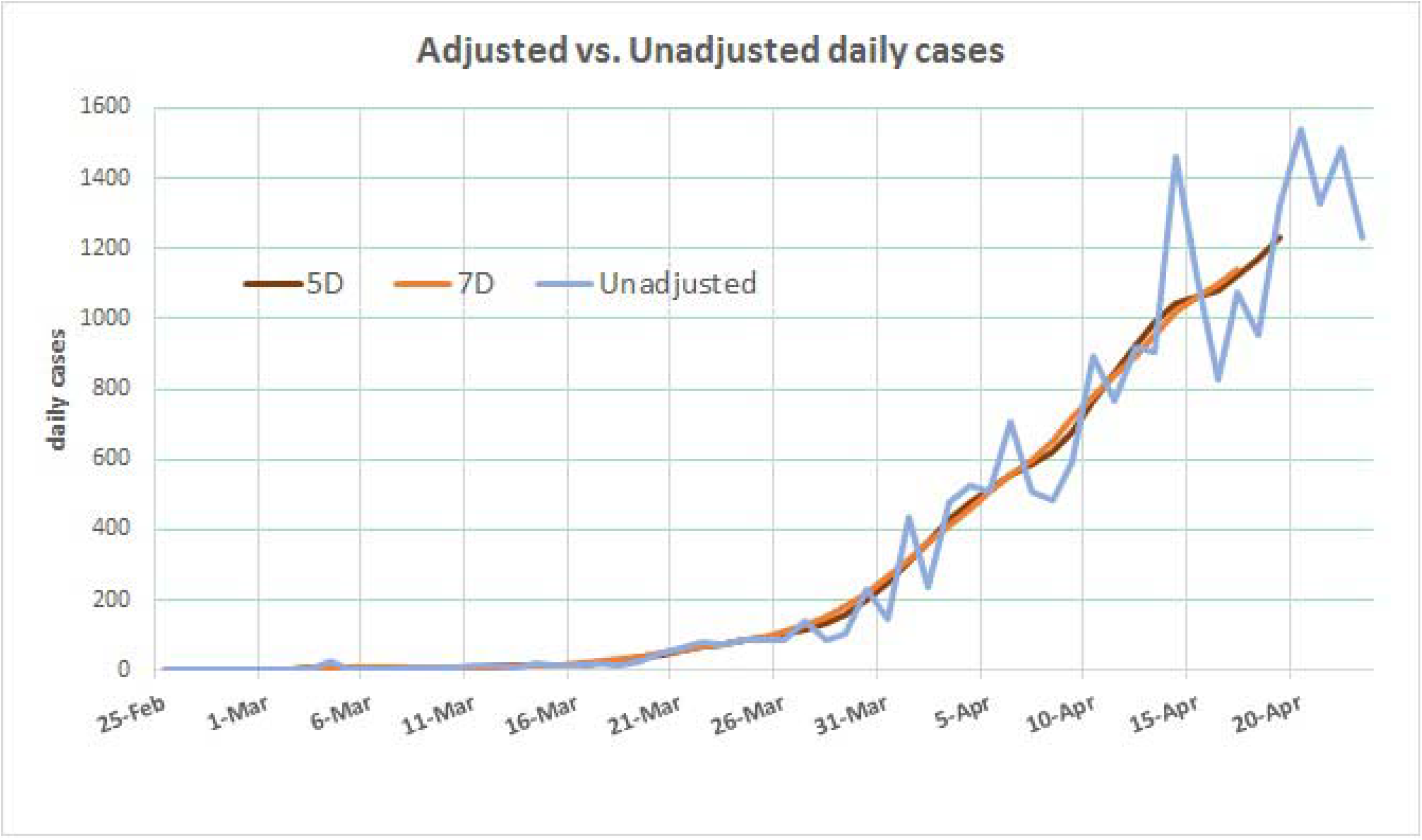
Plot of Unadjusted daily cases (*Udc*_*n*_) (up to 23 April) and Adjusted daily cases) *Adc*_*n*_ estimated from 5D cycle (up to 19 April) and 7D cycle (17 April).

### 3.5 Future course of Epidemic

ARIMA model forecasted (Fig. 6) formation of peak on 28 April for *AR*_*cn*_ of 5D cycle whereas for 7D cycle it forecasted a gradual decrease. Similarly, neural network model (NNM) forecasted (Fig. 7) towards formation of peak on 30 April for 5D cycle and gradual increase in *AR*_*cn*_ for 7D cycle.

**Figure 6.**
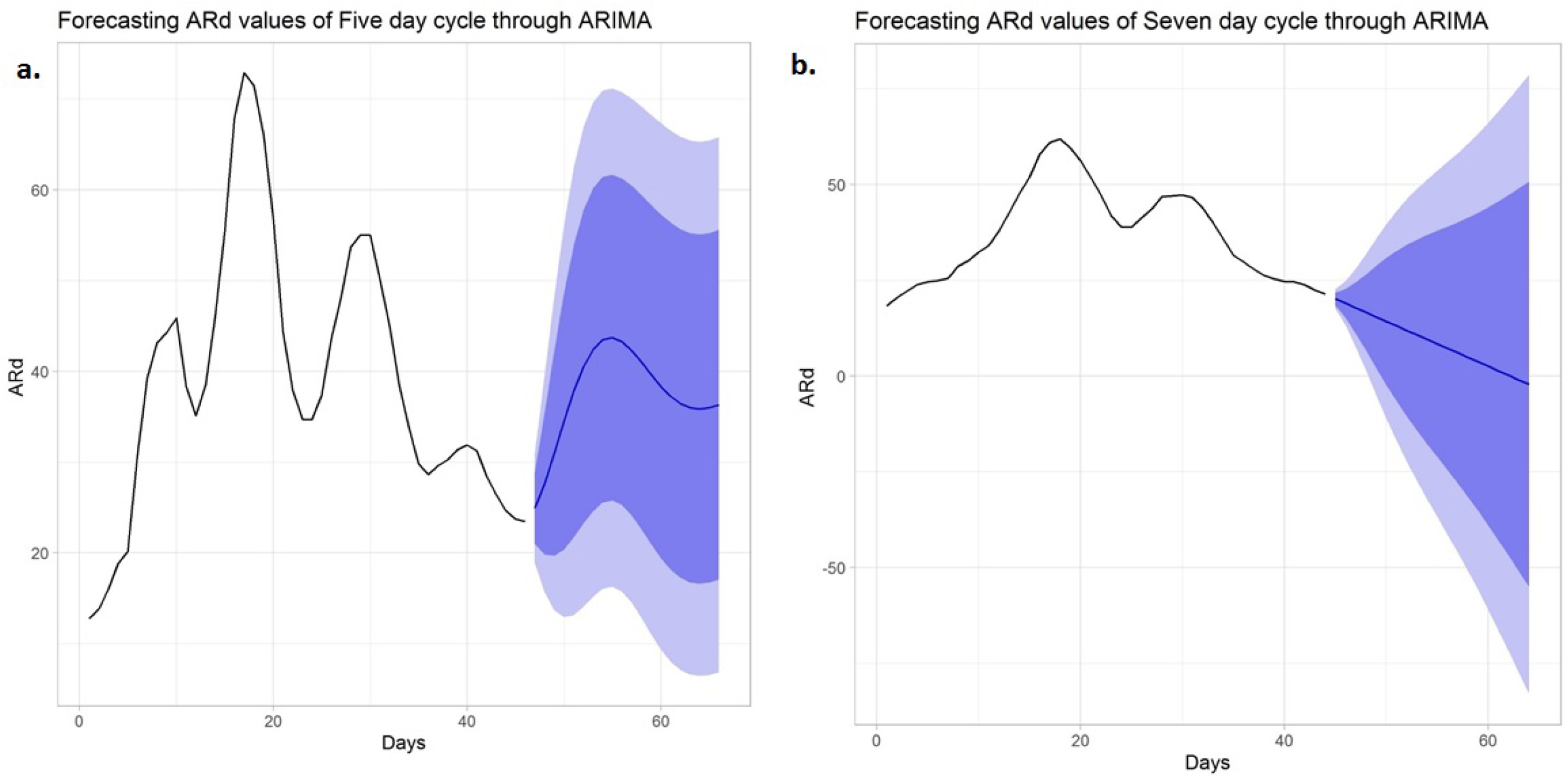
Forecasting *AR*_*cn*_ for five day cycle (5D) 20 March to 9 May and seven day cycle (7D) from 18 March to 7 May through ARIMA.

**Figure 7.**
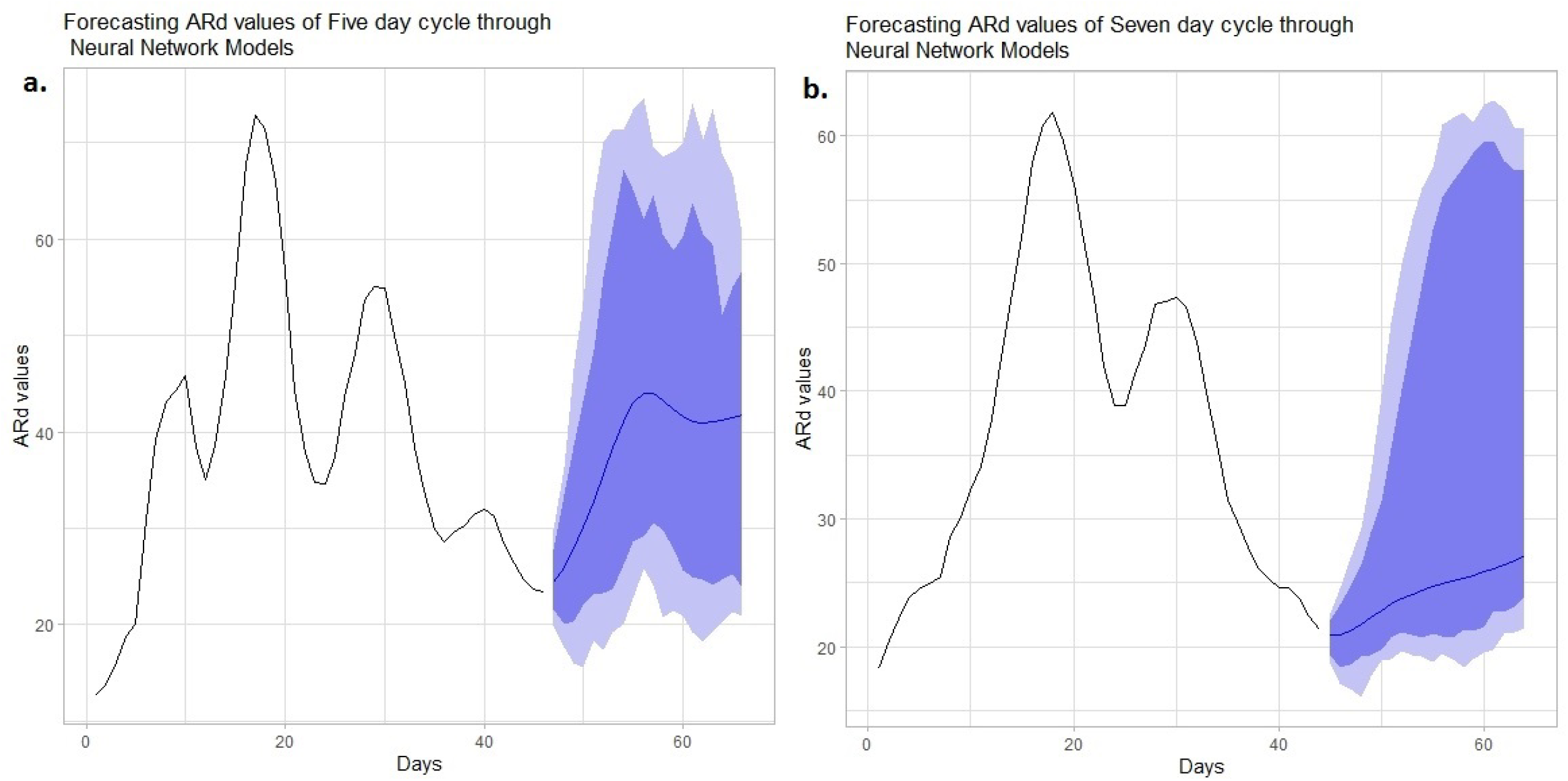
Forecasting *AR*_*cn*_ for five day cycle (5D) 20 March to 9 May and seven day cycle (7D) from 18 March to 7 May through Neural Network Models.

As seen from the *R*_*cmi*_ plot, though the rate of infection decreasing it the rate at which it is decreasing is slowing down. The last *R*_*cmi*_value for 7D cycleincreased from 128.22 (*R*_769_) to 129.69 (*R*_779_), similar trend was observed for 5D cycle which indicates a possible rise COVID-19 positive cases. Presently India is in growth stage where *Adc*_*n*_ is continuously rising. With recurrent peaks occurring, India may not reach critical point within May 3, the scheduled date to end the nationwide lockdown. Reducing *R*_*cmi*_ and *AR*_*cn*_ can be achieved through rigorous contact tracing and alternate testing methods such as Rapid antibody testing, CRISPAR (Liu et al., 2020) and sample pooling method (Hogan et al., 2020) can be useful for a large nation as India.

### 3.6 Limitations of the method

As of other statistical methods there are few limitations for the present method. When the spread of disease is slow the *AR*_*cn*_ and *R*_*cmi*_plot can’t depict the pattern of disease spread. Thus, the method is useful where local transmission or community spread of disease is prevalent. Pattern of disease spread is delineated by the method is based on reported data and doesn’t consider the underlaying unreported cases.

## 4. Conclusion

*AR*_*cn*_ and *R*_*cmi*_ plots help us in understanding the pattern of Epidemic and predicting the future scenario by minimizing the noise using in the raw data. The growth of epidemic in India has been non-uniform and is currently growing in linear fashion with *Adc*_*n*_ crossing 1,000. Lockdown combined with greater testing capacity is crucial to contain the spread of novel coronavirus. In the current study, we employed ingenious method to classify data into5D and 7D cycle to assess spread of virus and predict the future progression of epidemic. The method is simple and robust which uses publicly and most commonly available data.

## Data Availability

All the data was obtained from John Hopkins University GitHub repository.

https://github.com/CSSEGISandData/COVID-19

## Acknowledgements

The authors are thankful to all resource persons for their useful suggestions. No funding to declare.

